# Large-scale genetic analysis of age-related macular degeneration (AMD) drug targets highlights precision therapy opportunities for patients with high polygenic risk

**DOI:** 10.64898/2025.12.26.25343053

**Authors:** Jonathan S. Packer, Alice Zheng, Travis Mize, Kathleen D. Ferar, Lisa Bedford, Eimear E. Kenny, Krishna G. Aragam, Mary Helen Black

## Abstract

**Background:** In the past two decades, genetic studies have elucidated the contributions of key biological pathways to the pathogenesis of age-related macular degeneration (AMD), including complement activation, lipoprotein metabolism, angiogenesis, and extracellular matrix maintenance. Of these pathways, complement in particular has been observed to dominate the genetic architecture of AMD. Yet, clinical treatment of AMD with complement inhibitors has met with limited success.

**Methods:** Using data from four large-scale cohorts spanning 30,251 AMD cases and 438,016 AMD controls, we identified functional genetic variants to serve as proxies for complement inhibitor drug effects, and assessed their interaction with a pathway-specific AMD polygenic risk score (PRS). In each cohort, subjects were divided into low, medium, and high AMD risk groups based on quantiles of the PRS, such that each risk group included one-third of the cohort’s AMD cases. Drug target variant associations with AMD were evaluated in each risk group, as well as in all-comers. Quantitative biomarker analysis leveraging retinal phenotypes derived from optical coherence tomography (OCT) data was also performed.

**Results:** Genetic proxies for *C3* and *CFB* inhibition had an effect on AMD risk that was 1.6 to 2.3 times higher in the high PRS group compared to all-comers. Interactions between genetic drug proxies and the PRS was statistically significant, with replication across cohorts. Statistical support was strongest in three cohorts for *C3* and across all four cohorts for *CFB*. Examining a retinal thickness phenotype (ISOS-RPE), genetic drug proxy by PRS interaction was nominally significant for *CFB*, and directionally consistent for *C3*. Our results point to a continuous relationship between genetic complement activation / inhibition and AMD risk, across disease stages, without threshold effects.

**Conclusion:** Our findings suggest that patient heterogeneity due to genetically-influenced complement activation may explain the limited efficacy of AMD treatment with complement inhibitors to date. Prospective studies are warranted to assess whether precision therapy with complement inhibitors may be achieved by enrichment of patients with high PRS in future trials.

## Introduction

The first genome-wide association study (GWAS), published in 2005, reported the association of a missense variant in complement factor H (*CFH*) with risk of age-related macular degeneration (AMD).^1^ Subsequent GWAS have confirmed that variants contributing to dysregulation of the complement system — specifically, of the alternative complement pathway — represent a dominant feature of AMD’s genetic architecture.^2,3^ Complement pathway genes implicated by GWAS include complement components (*C3, C5, C9*); complement factors (*CFB, CFD, CFH, CFHR1, CFI*); *CD46*, a cofactor of *CFI* and negative regulator of complement activation^4–6^; vitronectin (*VTN*), an extracellular matrix component that inhibits formation of the membrane attack complex^7,8^; and *HTRA1*, a protease that cleaves vitronectin and fibromodulin, and triggers inflammation.^9–11^ Later GWAS also highlighted the contributions of non-complement loci, including *e*.*g*. genes associated with cholesterol and lipoprotein metabolism (*APOE, LIPC, CETP*), extracellular matrix maintenance (*TIMP3*), and angiogenesis (*VEGFA*).^12^

There are two main subtypes of AMD: wet and dry. Wet AMD, also called choroidal neovascularization (CNV), is characterized by abnormal blood vessels penetrating the barrier between the choroid and the retina, leading to fluid leakage, cell damage, and vision loss. Advanced dry AMD, known as geographic atrophy (GA), is characterized by RPE and photoreceptor cell death in the absence of neovascularization. Earlier stages of AMD disease progression (categorized as “dry”) can progress into either CNV or GA. “Early” / “intermediate” AMD is typically defined based on the size and abundance of drusen — yellow deposits, filled with proteins, lipids, and cellular debris, that form under the retina — as well as other retinal / pigmentary characteristics. GWAS effect sizes for CNV, GA, intermediate AMD, and early AMD are all highly correlated.^2,13^

Wet AMD is effectively treated with antagonists of vascular endothelial growth factor (VEGF), while GA is treated with complement inhibitors, which are only marginally effective at slowing disease progression. Pegcetacoplan, an intravitreally-delivered peptide C3 inhibitor, and avacincaptad pegol, and intravitreal RNA aptamer C5 inhibitor, were approved by the US FDA for treatment of GA in 2023. Both drugs slowed GA progression by roughly 20% over 1-to-2 years of follow-up.^14–16^ They were the first drugs approved for the GA subtype after previous failures in the indication, *e*.*g*. eculizumab, an intravenous C5 antibody, and lampalizumab, an intravitreal CFD antibody. Clinical trials in GA for additional complement inhibitors are ongoing, *e*.*g*. iptacopan, an oral small molecule CFB inhibitor (NCT05230537); vonaprument, an intravitreal C1q antibody fragment (NCT06510816); and pozelimab + cemdisiran, a combination of subcutaneous C5 antibody and subcutaneous liver-targeted C5 siRNA (NCT06541704).

Given the overwhelming genetic evidence for the centrality of the complement system in AMD pathogenesis, the underwhelming performance of complement inhibitors in GA is perplexing. It is possible that partial complement inhibition is insufficient to halt GA, although genetic variants in/near complement genes have been shown to have relatively large effects, even in a heterozygous state. It is also possible that earlier intervention is necessary for complement inhibitors to be effective in preventing disease progression. However, variants in complement pathway genes are associated with disease risk at every stage: early, intermediate, and advanced.^2,13^ GWAS-identified variant effect sizes across GA and CNV subtypes are remarkably correlated. Moreover, for any given variant, studies report greater effect size in advanced AMD compared to intermediate AMD. This suggests that these variants affect both risk of progression from no/early AMD to intermediate stages of disease as well as risk of progression from intermediate to advanced AMD. The genetic evidence is supported by observational studies: terminal complement activation (*i*.*e*. membrane attack complex formation and deposition) has been observed in the choroid and Bruch’s membrane of healthy subjects as young as 5 years old, and increases with age and AMD disease stage.^17^ As such, it is thought that complement activation in the retina may help clear cellular debris produced when retinal pigment epithelium phagocytoses and lyses photoreceptor outer segments.^17, 18^ Lastly, it is possible that varying response to treatment with complement inhibitors is largely explained by patient heterogeneity. We hypothesize that AMD pathogenesis is represented by a spectrum, ranging from disease driven by genetically-influenced complement hyperactivation to disease driven by complement-independent, non-heritable environmental factors. Biomarker data supports this hypothesis: while markers of complement activation are statistically significantly elevated in AMD cases vs. controls, case-control distributions overlap considerably.^19,20^

Thus, we sought to investigate the patient heterogeneity hypothesis from a genetic perspective, using pharmacomimetic (*i*.*e*. drug-mimicking) variants in *C3* and *CFB*. Specifically, we tested whether the effect of these variants on AMD risk is modified by background genetic predisposition to complement hyperactivation, as modeled by a complement pathway polygenic risk score (PRS).

## Methods

### Cohorts

We examined individual-level genetic and phenotypic data from four cohorts: UK Biobank (UKB), Electronic Medical Records and Genomics Network (eMERGE) III, Mass General Brigham Biobank (MGBB), and the International AMD Genetics Consortium (IAMDGC). UKB is a population-based biobank that broadly sampled ∼500,000 participants in the United Kingdom. eMERGE and MGBB employed hospital-based ascertainment of participants in the United States. IAMDGC is a case-control cohort in which the majority of participants were recruited from ophthalmology clinics across the United States, Europe, Australia, and Israel.

### Phenotype definitions

In UKB, MGBB, and eMERGE, we defined AMD cases as subjects who had at least one instance of ICD-10 H35.3 (“Degeneration of macula and posterior pole”) or ICD-9 362.5. Cases were further required to have no instances of ICD-10 H35.5 (“Hereditary retinal dystrophy”), H36.0 (“Diabetic retinopathy”) or ICD-9 362.0, 362.70-362.73, 362.75, 372.79. In UKB, we additionally classified any subject who had at least one instance of OPCS-4 C82 (“Destruction of lesion of retina”) or X93.1 (“Subfoveal choroidal neovascularisation drugs”), and/or who self-reported AMD at biobank enrollment (data field 20002 code 1528) as an AMD case. Across all cohorts, controls were defined as subjects who were not cases, and additionally did not have any instances of ICD-10 E10.3, E11.3, E12.3, E13.3, E14.3 (“Diabetes with ophthalmic complications”), H35 (“Other retinal disorders”), H36.0 (“Diabetic retinopathy”), H54.0 (“Blindness, binocular”), H54.1 (“Severe visual impairment, binocular”), H54.4 (“Blindness, monocular”), or ICD-9 362.0, 362.5, 362.70-73, 362.75, 362.79, 369.0-1, 369.6. For UKB, we expanded the control exclusion criteria to include OPCS-4 C79, C80, C81, C82, C83, C84, C85, X93 (ocular procedures and drug administration), and self-reported diabetic eye disease, retinitis pigmentosa, or macular degeneration (data field 20002 codes 1276, 1527, 1528 + data field 6148 codes 1, 5).

In UKB, we defined CNV cases as subjects who had at least one instance of OPCS-4 C82 or X93.1. We defined dry AMD cases as AMD cases who were not CNV cases. We used the any-AMD control set for analyses of CNV and dry AMD.

In IAMDGC, subjects were clinically assessed for CNV, GA, and intermediate AMD. Intermediate AMD was defined as the presence of pigmentary changes in the RPE or more than five macular drusen greater than 63 μm, requiring age at first diagnosis ≥ 50 years, in the absence of CNV or GA.

Quantitative biomarker analysis was conducted in UKB using retinal thickness measurements (category 100079, “Advanced boundary segmentation [TABS]”). These phenotypes were previously derived by Patel *et al*. and Ko *et al*. from optical coherence tomography (OCT) imaging performed in a subset (∼16%) of UKB subjects.^21,22^ Prior to statistical analysis, we excluded subjects who were <60 years old at the time of the OCT imaging, computed the average measurements from the left and right eyes, and applied a rank-based inverse normal transformation.

### Pharmacomimetic variants

We examined two pharmacomimetic missense variants in *C3*: 1) GRCh38-19-6718376-G-C (rs2230199) = *C3* R102G, which is also in strong linkage disequilibrium with *C3* P314L. 2) GRCh37-19-6718135-T-G (rs147859257) = *C3* K155Q.

*C3* K155Q results in resistance to C3 inactivation by *CFI* and *CFH*.^23^ To our knowledge, there is no published experimental characterization of *C3* R102G. However, manual inspection of supplementary tables from three proteomics publications showed a consistent association of R102G with reduced C3b levels and increased C3d levels, suggesting that the variant increases the rate of C3b to C3d conversion.^24–26^ Thus, both variants were considered to increase complement activation.

We examined four pharmacomimetic non-coding variants at the *CFB* locus: 1) GRCh38-6-31962685-G-A (rs429608, in moderate LD with *CFB* R32Q), 2) GRCh38-6-31979015-G-A (rs2746394, in strong LD with *SKIC2* R32W), 3) GRCh38-6-32187804-A-G (rs204993), and 4) GRCh38-6-31979250-G-T (rs181705462). These variants were previously reported as having conditionally independent, genome-wide significant associations with AMD.^2^ While there is not conclusive evidence that each of these *CFB* locus variants affects *CFB* expression or function, we note that GWAS associations of *C3, CFH*, and *CFI* with AMD implicate the alternative complement pathway in AMD pathogenesis, and outside of the *CFB-C2-C4A-C4B* locus, there are no other AMD GWAS loci that implicate the classical complement pathway. Second, *CFB* R74H, a rare coding variant, has been reported to have a strong protective effect against AMD.^27^ While *CFB* R74H is most often linked to another missense variant, *C2* P37L, a rare haplotype that has *CFB* 74H (minor allele) but *C2* 37P (major allele) is protective against AMD, indicating that the *CFB* 74H allele has an independent effect. Third, *CFB* R32Q, in modest LD in Europeans with the AMD GWAS variant rs429608, has been reported to reduce C3 to C3b conversion.^28^ Fourth, a Ph2 clinical trial of iptacopan is currently underway for the treatment of AMD (NCT05230537), following success in trials of IgA nephropathy, pointing to AMD as a plausible alternate indication benefitting from CFB inhibition.

### Pharmacomimetic scores

We aggregated the effects of the two *C3* variants listed above into a *C3* pharmacomimetic score. This score was defined as the weighted sum of the alternate allele dosages for each variant, where the weights were taken as variant-specific GWAS effect sizes from the Fritsche *et al*. IAMDGC joint model of 52 variants^2^. Similarly, we aggregated the four *CFB* variants listed above into a *CFB* pharmacomimetic score.

### Polygenic risk scores

We used the same weighted-sum method described in the previous section to define a complement pathway-specific AMD polygenic risk score (PRS). This PRS included 18 variants at the following loci: *C3, C9, CFB, CFH, CFI, VTN, HTRA1*.

For comparison, we also defined a genome-wide AMD PRS using 48 out of 52 variants reported by the Fritsche *et al*. IAMDGC study.^2^ We excluded 4 variants: rs3138141, rs121913059, and rs142450006 were not present in the UKB genotype data; rs191281603 was not significant in the IAMDGC joint model.

Due to overlap between individual-level IAMDGC data used for PRS training and a subset of interaction analyses presented, we re-weighted the variants using an inverse variance weighted meta-analysis of: 1) a joint model in UKB of the PRS variants’ association with AMD, and 2) GWAS effect sizes for each variant in the FinnGen r9 AMD GWAS.^29^ The meta-analysis weights were highly correlated with the original IAMDGC weights (R^2^ = 0.87).

The PRS variants and weights included in the present analyses are listed in Table S1.

We also compared PRS to single-gene scores for *CFH* and *HTRA1*. These were constructed using the method described above, with variants annotated as residing in/near the *CFH* or *HTRA1* loci in Fritsche *et al*.^*2*^

### Statistical analysis of individual-level data

We restricted all analyses to unrelated subjects of genetically-defined European ancestry. Sample sizes for non-European subjects, in the available individual-level data, were small (Table S2, *e*.*g*. < 550 African AMD cases) and insufficient for the genetic interaction analyses that are the central focus of our study. Interaction tests require larger sample sizes compared to single-variant association tests. In subjects of African genetic ancestry, AMD GWAS effect sizes at complement gene loci are substantially attenuated, further reducing statistical power.^3^

We used logistic regression models to assess the association of the pharmacomimetic (PM) scores, the AMD PRS, and the PM scores in interaction with the PRS with risk of AMD. In each model, we included the following covariates: age, age squared, sex, age^*^sex, age^2*^sex, and cohort-specific principal components of genetic ancestry. We defined “age” as “age at right censoring”, *i*.*e*. a subject’s age on the date of the most recent phenotypic data in their cohort (considering all subjects in the cohort), or the subject’s age at death, whichever came first.

Before fitting any regression model that included an interaction term for a PM score and an AMD PRS, we first adjusted the PRS by linear regression to remove the contribution of the variants in the PM score to the PRS. We confirmed that this *post-hoc* adjustment of the PRS gave identical results to entirely re-constructing the PRS excluding the PM variants.

AMD cases and controls in each cohort were stratified into three bins, corresponding to low, medium, and high AMD risk, based on values of the AMD complement pathway PRS. PRS cut-offs were selected within each cohort such that each bin captured one-third of the AMD cases. In several analyses, we evaluated the ratio of the model coefficient representing a PM score’s effect on AMD risk in subjects in the high PRS bin, to the model coefficient representing that PM score’s effect on AMD risk in the full cohort.

Additional imaging biomarker analyses were performed using the quantitative retinal thickness measures described above with linear regression models adjusted for the same covariates (age, age squared, sex, age^*^sex, age2^*^sex, and cohort-specific principal components of genetic ancestry).

### Meta-analysis of AMD GWAS summary statistics

To provide further context on the 18 variants in the AMD complement pathway PRS, we compared the associations of each variant with overall AMD risk in a GWAS meta-analysis to the corresponding associations of each variant in analyses of stage-specific AMD phenotypes.

We used METAL^30^ to perform an inverse variance weighted meta-analysis of European-ancestry AMD GWAS summary statistics from UK Biobank, FinnGen r11^29^, MVP^31^, and GERA.^32^ For each study except UKB, published GWAS summary statistics were used without further processing. For UKB, we performed GWAS using REGENIE^33^ to test variant association with the binary AMD phenotype described above, adjusted for the same covariates (age, age squared, sex, age^*^sex, age2^*^sex, and cohort-specific principal components of genetic ancestry). We assessed variant associations with early AMD using GWAS summary statistics from Winkler *et al*.^*13*^. We assessed variant associations with intermediate AMD, advanced AMD, and “advanced AMD conditional on intermediate AMD” by applying logistic regression models to individual-level data from the IAMDGC cohort (described above).

## Results

Altogether, the four cohorts we investigated included 30,251 AMD cases and 438,016 AMD controls (Table 1, Table S3). In each cohort, the *C3* and *CFB* pharmacomimetic scores, the 18-variant AMD complement pathway PRS, and the 48-variant AMD genomewide PRS were predictive of AMD case-control status (Table 2). Complement pathway PRS effect sizes were similar to those of the genomewide PRS across the 4 cohorts.

**Table 1.** Cohorts used in the analysis. The “AMD subtypes” column refers to which disease subtypes could be discerned in a given cohort using the available phenotype information. CNV = choroidal neovascularization, GA = geographic atrophy. In the UK Biobank, Dry AMD was defined as any AMD that could not be determined to be CNV.

| Cohort | Cohort type | AMD cases | AMD controls | AMD subtypes available for analysis |
| --- | --- | --- | --- | --- |
| UK Biobank | Population-based biobank | 9,638 | 331,639 | Dry, CNV |
| IAMDGC | Clinical cohort | 14,075 | 12,054 | Intermediate, GA, CNV |
| eMERGE | Hospital biobanks | 4,117 | 56,328 | (none) |
| MGBB | Hospital biobank | 2,421 | 37,995 | (none) |

**Table 2.** Associations of the genetic instruments used in the analysis with AMD case-control status, in each cohort. PM = pharmacomimetic. PRS = polygenic risk score. OR = odds ratio. IAMDGC has the largest odds ratios, likely because the AMD phenotyping in this clinical cohort was more precise than in the other cohorts (in which AMD was identified from ICD codes).

| Genetic instrument | UKB | IAMDGC | eMERGE | MGBB |
| --- | --- | --- | --- | --- |
| C3 PM score | OR = 1.09<br>$p = 1.8 \times 10^{-18}$ | OR = 1.26<br>$p = 2.9 \times 10^{-62}$ | OR = 1.07<br>$p = 2.8 \times 10^{-4}$ | OR = 1.05<br>$p = 3.4 \times 10^{-2}$ |
| CFB PM score | OR = 1.08<br>$p = 1.3 \times 10^{-11}$ | OR = 1.29<br>$p = 9.1 \times 10^{-82}$ | OR = 1.11<br>$p = 7.6 \times 10^{-8}$ | OR = 1.10<br>$p = 5.0 \times 10^{-5}$ |
| AMD complement PRS | OR = 1.39<br>$p = 7.1 \times 10^{-226}$ | OR = 2.43<br>$p < 10^{-300}$ | OR = 1.39<br>$p = 6.3 \times 10^{-66}$ | OR = 1.32<br>$p = 2.9 \times 10^{-38}$ |
| AMD genomewide PRS | OR = 1.41<br>$p = 4.4 \times 10^{-236}$ | OR = 2.58<br>$p < 10^{-300}$ | OR = 1.41<br>$p = 1.6 \times 10^{-73}$ | OR = 1.33<br>$p = 2.8 \times 10^{-39}$ |

In individuals with high complement pathway PRS, the *C3* and *CFB* scores were substantially more predictive of AMD case-control status than in those with medium or low PRS (Fig 1A). This pattern was consistent across each of the four cohorts evaluated (*C3* score x PRS model *p*_interaction_ = 2.9 x 10^-12^ in UKB, 1.5 x 10^-5^ in IAMDGC, 1.5 x 10^-5^ in eMERGE, 0.17 in MGBB; *CFB* score x PRS model *p*_interaction_ = 8.3 x 10^-7^, 3.7 x 10^-8^, 0.036, 0.009). The ratio of the *C3* and *CFB* score effects in the high PRS group vs. all-comers was also similar across cohorts, ranging from approximately 1.6 to 2.3 (ratios for the *C3* score: 1.84 in UKB, 1.56 in IAMDGC, 2.31 in eMERGE, and 1.82 in MGBB; ratios for the *CFB* score: 1.92, 1.57, 1.83, and 1.86). The pattern of PRS stratification of *C3* and *CFB* score effects was similar when dry AMD was modeled as the disease outcome instead of any-AMD, in cohorts for which dry and wet subtypes of AMD were able to be distinguished (Fig 1B).

**Figure 1:**
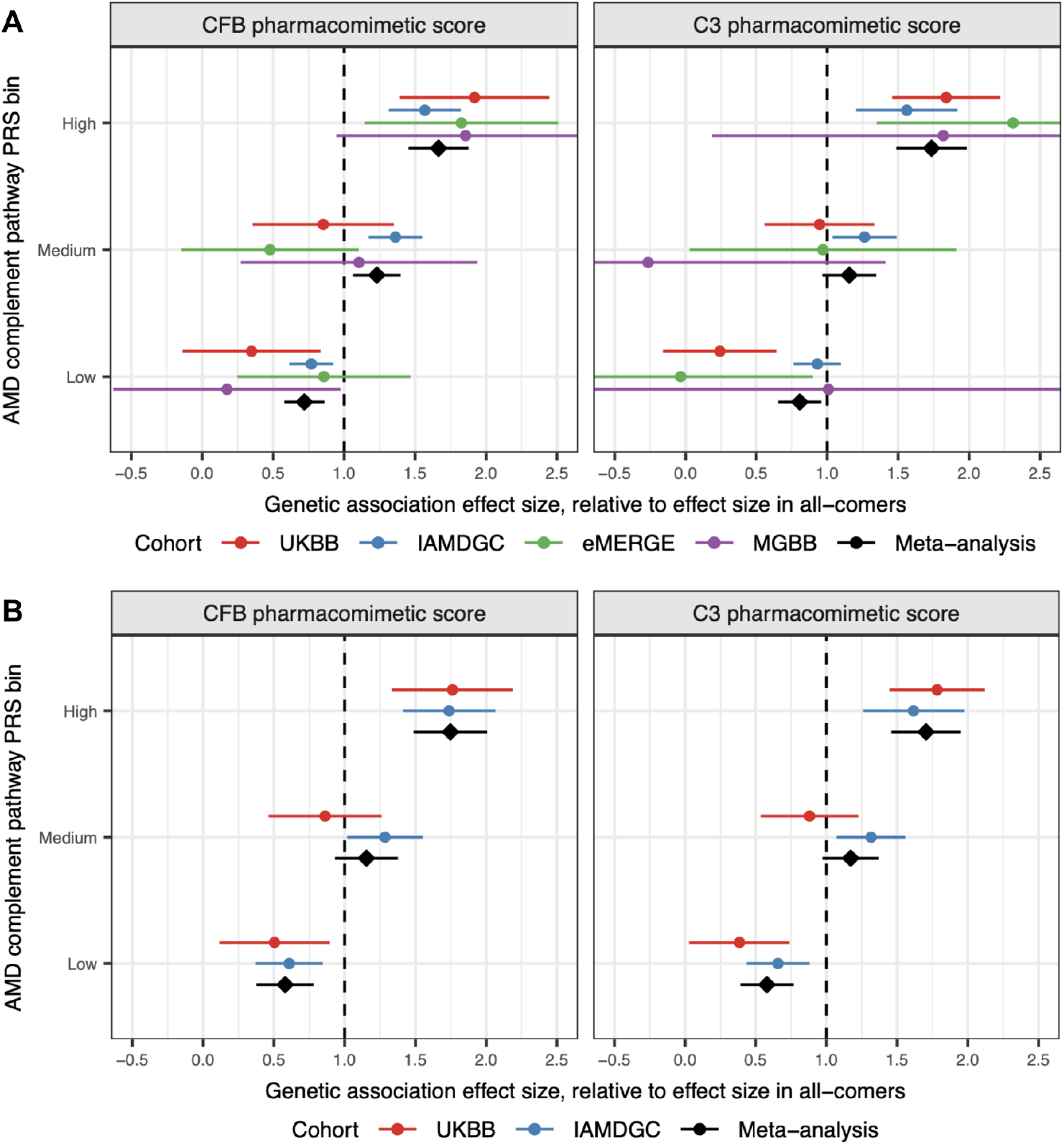
**A**. Effects of the *CFB* and *C3* pharmacomimetic scores (units = standard deviations) on risk of any AMD, in individuals defined as having low, medium, or high AMD complement pathway PRS. The PRS cut-offs for the three subgroups were selected such that each subgroup represents exactly one-third of the AMD cases within a given cohort. **B**. As panel A, but with dry AMD assessed as the outcome.

Two loci, *CFH* and *HTRA1*, account for a disproportionately large share of the variance in AMD liability that is explained by common genetic variants.^2^ We evaluated digenic models, in which the pharmacomimetic scores for *C3* and *CFB* were tested for interaction with single-gene risk scores for *CFH* and *HTRA1*, and compared them to our previous polygenic model (Table S4). There was strong evidence for a digenic *CFB*-*CFH* interaction (*p* < 0.05 in all 4 cohorts) and *C3-CFH* interaction (*p* < 0.05 in 3 cohorts). The ratio of the *CFB*-*CFH* interaction effect size to the *CFB*-PRS interaction effect size was 70% in UKB, 82% in IAMDGC, and >100% in eMERGE and MGBB. For *C3*, the corresponding statistics were 70%, 67%, 80%, and 19%. Taken together, our findings suggest the complement pathway-based PRS is a more effective stratifier of *C3*- and *CFB*-specific effects, and therefore response to treatment with complement inhibition.

*C3* and *CFB* score interactions with AMD complement pathway PRS were recapitulated in retina optical coherence tomography (OCT) data from ∼45,000 UKB subjects. We tested each OCT-derived phenotype available (40 phenotypes) for association with dry AMD case-control status (Table S5). ISOS-RPE thickness in the central subfield, *i*.*e*. the thickness of the retina as measured from the junction of the inner and outer photoreceptor segments to the retinal pigment epithelium, yielded the strongest association. Reduced central subfield ISOS-RPE thickness was correlated with increased risk of dry AMD (Fig 2A). Correspondingly, the AMD risk-increasing *C3* and *CFB* scores correlated with reduced central subfield ISOS-RPE thickness, and the magnitude of this correlation was amplified in subjects with high AMD complement pathway PRS (Fig 2B).

**Figure 2:**
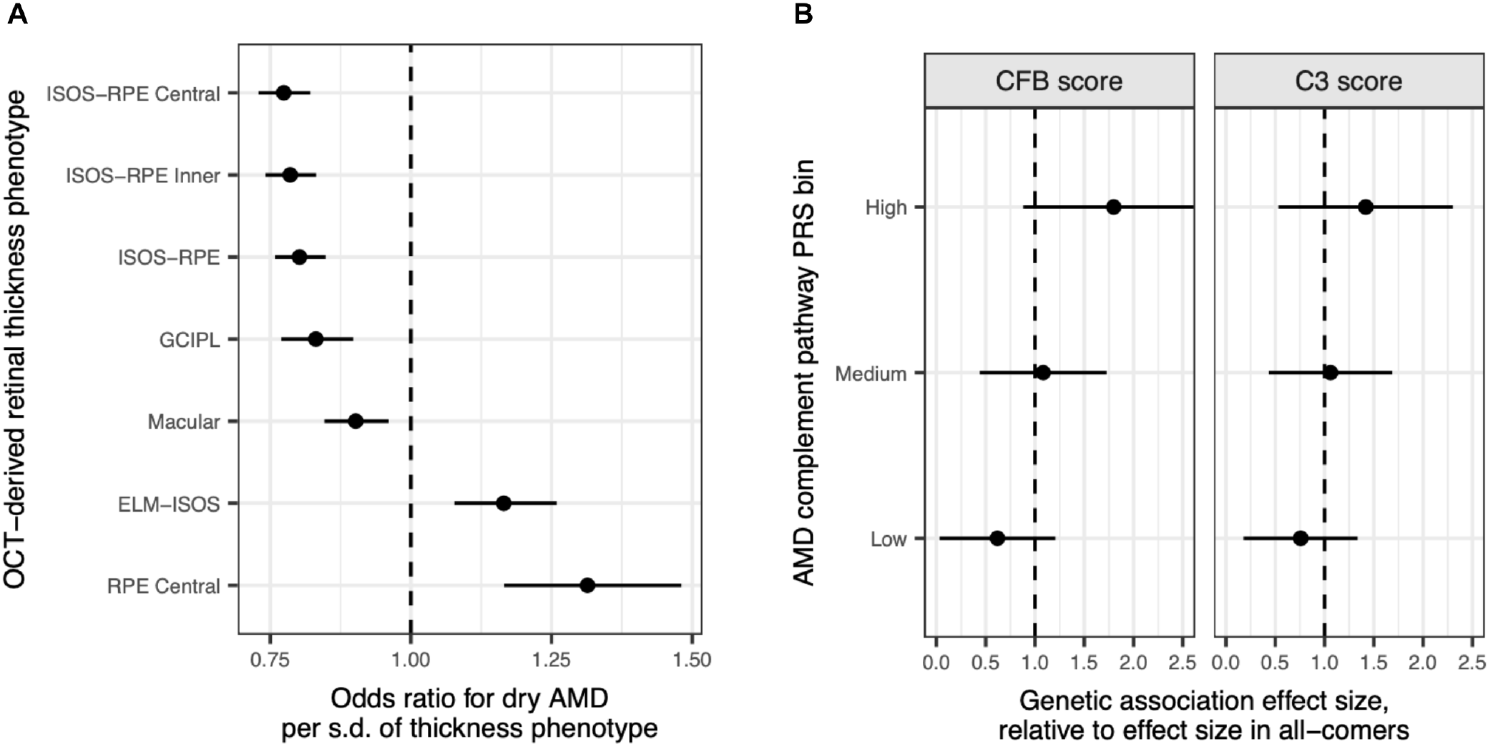
**A**. Association of selected retinal thickness phenotypes with dry AMD. The phenotypes were derived from UKB optical coherence tomography (OCT) data. Subjects with age <60 at time of OCT measurement were excluded. **B**. Effects of the *CFB* and *C3* pharmacomimetic scores on ISOS-RPE thickness in the central subfield, in groups of subjects defined as having low, medium, or high AMD complement pathway PRS. *CFB* score x PRS interaction is nominally significant (*p* = 0.014); *C3* score x PRS interaction is directionally consistent (*p* = 0.25).

Our results are consistent with the hypothesis that there is substantial heterogeneity across AMD patients represented by the extent to which the complement pathway is driving disease progression. For reporting of results (*cf*. Fig 1, Fig 2) this heterogeneity is represented by low, medium, and high PRS bins, but we note that the relationship between the AMD complement pathway PRS and log odds of AMD is approximately linear and continuous (Fig 3A). This indicates that 1) the distribution of common genetic variants results in a continuous spectrum in which individuals can have a low, medium, or high level of predisposition to complement hyperactivation; 2) an individual’s location on this spectrum is physiologically relevant to AMD; and 3) there is no genetic evidence for the existence of a critical complement inhibition threshold, within this physiologically-relevant range. Lastly, the observed effects of genetic variants on risk of “any AMD” are highly correlated with those at particular disease stages, *i*.*e*. early AMD, intermediate AMD, or advanced AMD, and with their effects on the likelihood of a patient having advanced AMD conditional on that patient having intermediate AMD (Fig 3B). This suggests that the contribution of genetically-regulated complement dysregulation to AMD is continuous throughout the disease course.

**Figure 3.**
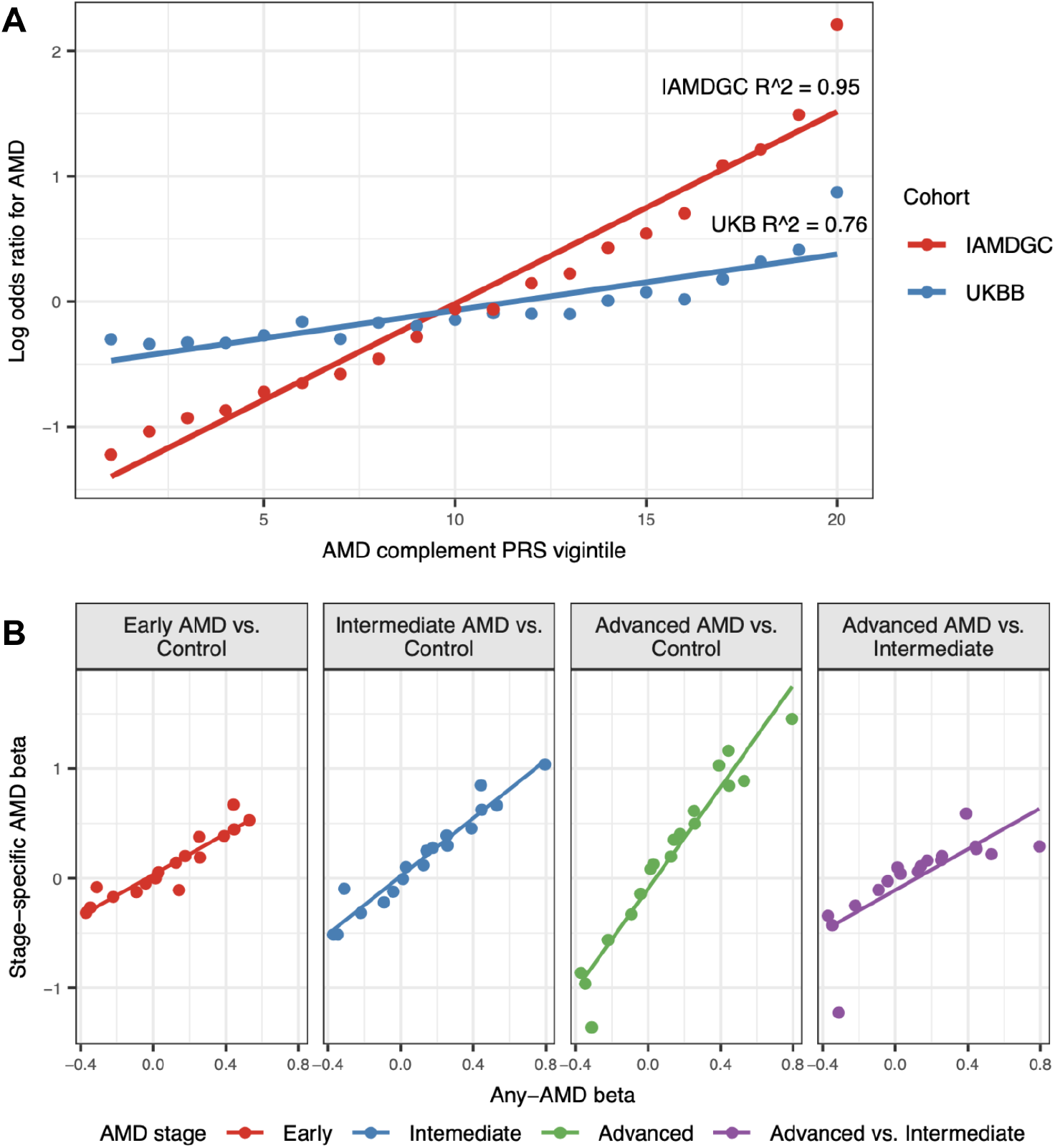
**A**. Correlation of AMD complement pathway PRS vigintiles (5%-iles) with the log odds of a subject having AMD, relative to all-comers in each respective cohort. Specifically, the log odds of a subject having AMD was computed for each PRS vigintile and divided by the log odds of a subject having AMD among those in the cohort (all-comers), irrespective of PRS. Correlation between the log odds ratio statistics and PRS vigintile number (1 to 20) was computed. The association of the PRS with AMD is stronger (*i*.*e*. slope is greater) in IAMDGC vs. UKB. This may be attributed to IAMDGC having clinically-assessed AMD, which is likely more accurate than the ICD-9/10 code-derived AMD extracted from UKB, especially in distinguishing *bona fide* controls for subjects with asymptomatic early / intermediate AMD. **B**. Associations of the 18 variants of the AMD complement pathway PRS with “any AMD”, compared to associations with stage-specific AMD phenotypes. Associations are presented as “betas”, *i*.*e*. the coefficient for the variant in a logistic regression model associating variant genotype with AMD phenotype. “Any AMD” associations were computed in a European-ancestry AMD GWAS meta-analysis (see Methods). Stage-specific AMD association statistics were retrieved from IAMDGC studies.^2,13^ The linear regression R^2^ statistics for the comparisons of the any-AMD betas to the stage-specific betas are as follows: 0.86 (early), 0.94 (intermediate), 0.94 (advanced), 0.60 (advanced vs. intermediate). The outlier variant in the advanced vs. intermediate analysis (right-most panel) is *CFH* rs148553336. Excluding this variant, the R^2^ is 0.76. One variant, *CFI* rs141853578, was missing from the early AMD GWAS^13^ and was thus excluded from the early AMD analysis (left-most panel).

## Discussion

Given the underwhelming success of complement inhibition in the treatment of AMD despite the strength of genetic evidence for the role of complement activation in disease pathogenesis, we hypothesized genetic heterogeneity to account for this disparity and conducted analyses of four large-scale human cohorts spanning nearly 470,000 individuals to test this hypothesis. Our findings indicate that polygenic predisposition to complement hyperactivation in AMD, represented by a novel complement pathway-specific PRS, modifies the magnitude of genetically inferred *C3/CFB* effects on AMD risk, supporting the hypothesis that complement inhibition may yield larger benefit in genetically high-risk individuals.

Considering the modest performance of complement inhibitors in GA patients to date, the large number of poorly differentiated complement inhibitors currently in preclinical and clinical stages of development, and the persistently high unmet clinical need in this stage of AMD, these findings warrant further clinical investigation. Yet to our knowledge, only one emerging biopharma company (Character Bio, Series B announced March 2025)^34^ has publicly disclosed plans to develop a proprietary complement pathway PRS for use in phase 2 trials of CTX114, a novel complement inhibitor designed to delay or prevent retinal cell death and vision loss in patients with GA.^35^

Past clinical trials of complement inhibitors in geographic atrophy have used change in square-root transformed lesion area as their primary endpoint, as quantified by fundus autofluorescence (FAF). This endpoint is challenging, and affected by parameters that can be difficult to model, such as pre-treatment lesion size and growth rate. Accordingly, OCT-derived phenotypes have been investigated as potential endpoints.^36^ Our analysis of UK Biobank data demonstrates that genetic validation of AMD drug targets and patient stratification approaches can be recapitulated in OCT data, further supporting the arguments that have previously been made for using OCT in clinical development, especially for high-risk intermediate AMD.

Early-stage research into approaches for patient stratification has greatly outpaced clinical implementation, particularly for indications outside of oncology. Detailed discussion of this discordance is outside the scope of this paper. But, as we are considering patient stratification in AMD, we note one example of an FDA-approved therapy with a precision approach: the use of eosinophil count as a patient stratifier in the development of dupilumab, an antibody against IL4R which is used to suppress Th2 / eosinophil-mediated inflammation. In the United States, dupilumab is approved for asthma and chronic obstructive pulmonary disease characterized by a high eosinophil phenotype (eg. >300 cells/µL). We propose that the mechanistic link between eosinophil activity and dupilumab efficacy is analogous to that between our proposed stratifier (a PRS representing complement pathway activity) and the therapies to which it would be relevant (complement inhibitors).

A limitation of our study was that we were precluded from assessing statistical genetic interactions in individuals of non-European ancestry, as we had insufficient individual-level data available (*e*.*g*. fewer than 550 African ancestry AMD cases). Further investigation in diverse cohorts such as Million Veteran’s Program (MVP) and AllOfUs is warranted, particularly given the attenuation of complement variant associations with AMD in individuals who have local African ancestry at the complement gene loci.^3^ We also acknowledge that our analyses and insights are based on genetic proxies for drug effects, rather than treated trial participants. As such, they do not directly estimate drug efficacy and should be interpreted as hypothesis-generating for PRS-guided trial design.

In conclusion, our findings suggest that patient heterogeneity with respect to genetically-influenced complement activation may explain the limited efficacy of complement inhibitors in the treatment of AMD. Specifically, we observed substantially stronger genetic effects corresponding to complement inhibition among individuals with high complement pathway PRS, and this was true regardless of AMD subtype or disease stage. Prospective studies are needed to assess whether precision therapy targeting patients most likely to benefit from complement inhibitor treatment may be achieved by enrichment of high PRS patients in future clinical trials.

## Data Availability

All data analyzed in the present study are either freely publicly available, or available by direct application to UKB, dbGaP, or other sources.

