## Supplementary material for "Large-scale genetic analysis of age-related macular degeneration (AMD) drug targets highlights precision therapy opportunities for patients with high polygenic risk": Table S1

| Rsid | Chromosome | Position (GRCh37) | Position (GRCh38) | Effect allele | Other allele | Weight from IAMDGC | Alternate weight (UKB + FG meta-analysis) | Included in the complement pathway PRS? |
|---|---|---|---|---|---|---|---|---|
| rs10922109 | 1 | 196704632 | 196735502 | A | C | -0.673345 | -0.420778 | Yes |
| rs570618 | 1 | 196657064 | 196687934 | G | T | -0.553885 | -0.416811 | Yes |
| rs148553336 | 1 | 196613173 | 196644043 | C | T | -1.171183 | -0.381499 | Yes |
| rs187328863 | 1 | 196380158 | 196411028 | T | C | 0.385262 | 0.163055 | Yes |
| rs61818925 | 1 | 196815450 | 196846320 | G | T | -0.165514 | -0.000975 | Yes |
| rs35292876 | 1 | 196706642 | 196737512 | T | C | 0.431782 | 0.345755 | Yes |
| rs11884770 | 2 | 228086920 | 227222204 | C | T | 0.083382 | 0.047770 | No |
| rs62247658 | 3 | 64715155 | 64729479 | T | C | -0.131028 | -0.059079 | No |
| rs140647181 | 3 | 99180668 | 99461824 | C | T | 0.615186 | 0.090629 | No |
| rs55975637 | 3 | 99419853 | 99701009 | A | G | 0.148420 | 0.012279 | No |
| rs10033900 | 4 | 110659067 | 109737911 | C | T | -0.139762 | -0.065749 | Yes |
| rs141853578 | 4 | 110685820 | 109764664 | T | C | 1.633154 | 0.894665 | Yes |
| rs62358361 | 5 | 39327888 | 39327786 | T | G | 0.512824 | 0.230353 | Yes |
| rs114092250 | 5 | 35494448 | 35494346 | A | G | -0.342490 | -0.106805 | No |
| rs429608 | 6 | 31930462 | 31962685 | A | G | -0.673345 | -0.246150 | Yes |
| rs2746394 | 6 | 31946792 | 31979015 | A | G | 1.026042 | 0.168471 | Yes |
| rs204993 | 6 | 32155581 | 32187804 | G | A | 0.122218 | 0.039074 | Yes |
| rs181705462 | 6 | 31947027 | 31979250 | T | G | 0.444686 | 0.170610 | Yes |
| rs943080 | 6 | 43826627 | 43858890 | T | C | 0.139262 | 0.062338 | No |
| rs1142 | 7 | 104756326 | 105115879 | T | C | 0.131028 | 0.050211 | No |
| rs7803454 | 7 | 99991548 | 100393925 | T | C | 0.139762 | 0.018566 | No |
| rs13278062 | 8 | 23082971 | 23225458 | T | G | 0.116534 | 0.041985 | No |
| rs10781182 | 9 | 76617720 | 74002804 | G | T | -0.113329 | 0.013125 | No |
| rs71507014 | 9 | 73438605 | 70823689 | GC | G | -0.104360 | -0.033246 | No |
| rs1626340 | 9 | 101923372 | 99161090 | A | G | -0.127833 | -0.042789 | No |
| rs2740488 | 9 | 107661742 | 104899461 | C | A | -0.116534 | -0.045881 | No |
| rs12357257 | 10 | 24999593 | 24710664 | A | G | 0.113329 | 0.024813 | No |
| rs3750846 | 10 | 124215565 | 122456049 | C | T | 1.075002 | 0.511937 | Yes |
| rs61941274 | 12 | 112132610 | 111694806 | A | G | 0.470004 | 0.022039 | No |
| rs9564692 | 13 | 31821240 | 31247103 | T | C | -0.105361 | -0.043761 | No |
| rs61985136 | 14 | 68769199 | 68302482 | T | C | 0.127833 | 0.039875 | No |
| rs2842339 | 14 | 68986999 | 68520282 | A | G | -0.165514 | -0.056379 | No |
| rs2043085 | 15 | 58680954 | 58388755 | C | T | 0.139762 | 0.066814 | No |
| rs2070895 | 15 | 58723939 | 58431740 | A | G | -0.150823 | -0.070813 | No |

| | | | | | | | | |
|---|---|---|---|---|---|---|---|---|
| rs5817082 | 16 | 56997349 | 56963437 | CA | C | -0.139262 | -0.045198 | No |
| rs17231506 | 16 | 56994528 | 56960616 | T | C | 0.104360 | 0.076125 | No |
| rs72802342 | 16 | 75234872 | 75200974 | A | C | -0.235722 | -0.107725 | No |
| rs11080055 | 17 | 26649724 | 28322698 | C | A | 0.083382 | 0.031258 | Yes |
| rs6565597 | 17 | 79526821 | 81559795 | T | C | 0.113329 | -0.000211 | No |
| rs2230199 | 19 | 6718387 | 6718376 | C | G | 0.385262 | 0.173471 | Yes |
| rs147859257 | 19 | 6718146 | 6718135 | G | T | 1.169381 | 0.503553 | Yes |
| rs12019136 | 19 | 5835677 | 5835666 | A | G | -0.301105 | -0.164873 | Yes |
| rs67538026 | 19 | 1031438 | 1031439 | T | C | -0.105361 | -0.009908 | No |
| rs429358 | 19 | 45411941 | 44908684 | C | T | -0.400478 | -0.148457 | No |
| rs73036519 | 19 | 45748362 | 45245104 | C | G | -0.094311 | -0.026142 | No |
| rs201459901 | 20 | 56653724 | 58078668 | TA | T | -0.274437 | -0.010476 | No |
| rs5754227 | 22 | 33105817 | 32709831 | C | T | -0.235722 | -0.089050 | No |
| rs8135665 | 22 | 38476276 | 38080269 | T | C | 0.131028 | 0.023927 | No |
