## Supplementary material for "Large-scale genetic analysis of age-related macular degeneration (AMD) drug targets highlights precision therapy opportunities for patients with high polygenic risk": Table S2

| Ancestry | Cohort | AMD Cases | AMD Controls | Neither-case-nor-controls | AMD Prevalence |
|---|---|---|---|---|---|
| European (EUR) | UKBB | 9,638 | 331,639 | 15,650 | 2.7% |
| European (EUR) | IAMDGC | 14,075 | 12,054 | 0 | 53.9% |
| European (EUR) | eMERGE | 4,117 | 56,328 | 4,819 | 6.3% |
| European (EUR) | MGBB | 2,405 | 37,866 | 2,342 | 5.6% |
| European (EUR) | Total | 30,235 | 437,887 | 22,811 | NA |
| African (AFR) | UKBB | 115 | 5,647 | 481 | 1.8% |
| African (AFR) | IAMDGC | 0 | 0 | 0 | NA |
| African (AFR) | eMERGE | 242 | 11,806 | 1,495 | 1.8% |
| African (AFR) | MGBB | 171 | 2,236 | 350 | 6.2% |
| African (AFR) | Total | 528 | 19,689 | 2,326 | NA |
| Admixed American (AMR) | UKBB | 11 | 908 | 37 | 1.2% |
| Admixed American (AMR) | IAMDGC | 0 | 0 | 0 | NA |
| Admixed American (AMR) | eMERGE | 82 | 2,912 | 460 | 2.4% |
| Admixed American (AMR) | MGBB | 132 | 3,208 | 255 | 3.7% |
| Admixed American (AMR) | Total | 225 | 7,028 | 752 | NA |
| South Asian (SAS) | UKBB | 179 | 7,325 | 816 | 2.2% |
| South Asian (SAS) | IAMDGC | 0 | 0 | 0 | NA |
| South Asian (SAS) | eMERGE | 5 | 464 | 41 | 1.0% |
| South Asian (SAS) | MGBB | 22 | 520 | 42 | 3.8% |
| South Asian (SAS) | Total | 206 | 8,309 | 899 | NA |
| East Asian (EAS) | UKBB | 65 | 2,488 | 125 | 2.4% |
| East Asian (EAS) | IAMDGC | 0 | 0 | 0 | NA |
| East Asian (EAS) | eMERGE | 20 | 721 | 64 | 2.5% |
| East Asian (EAS) | MGBB | 44 | 941 | 60 | 4.2% |
| East Asian (EAS) | Total | 129 | 4,150 | 249 | 2.8% |
