## Supplementary material for "Large-scale genetic analysis of age-related macular degeneration (AMD) drug targets highlights precision therapy opportunities for patients with high polygenic risk": Table S3

| Statistic | Cohort | AMD cases (high complement PRS) | AMD cases (non-high complement PRS) | AMD Controls | All Subjects | All subjects with high complement PRS | All subjects with non-high complement PRS |
|---|---|---|---|---|---|---|---|
| Sample size | UKBB | 3,214 | 6,424 | 331,639 | 356,927 | 76,574 | 280,353 |
| Sample size | eMERGE | 1,374 | 2,743 | 56,328 | 65,264 | 16,388 | 48,876 |
| Sample size | MGBB | 807 | 1,614 | 37,995 | 40,416 | 9,685 | 30,732 |
| Sample size | IAMDGC | 4,693 | 9,382 | 12,054 | 26,129 | 5,729 | 20,400 |
| Mean age | UKBB | 76.8 | 75.9 | 70.4 | 70.7 | 70.7 | 70.7 |
| Mean age | eMERGE | 80.3 | 79.7 | 58.0 | 59.9 | 59.8 | 59.9 |
| Mean age | MGBB | 76.6 | 75 | 60 | 60.9 | 61.0 | 60.9 |
| Mean age | IAMDGC | 75.3 | 76.7 | 69.8 | 73.2 | 74.2 | 73.0 |
| % male | UKBB | 40.8% | 43.2% | 46.0% | 46.2% | 46.3% | 46.2% |
| % male | eMERGE | 44.1% | 43.3% | 46.8% | 47.4% | 46.7% | 47.6% |
| % male | MGBB | 48.3% | 47.8% | 45.0% | 45.2% | 45.0% | 45.3% |
| % male | IAMDGC | 40.0% | 39.3% | 43.0% | 41.1% | 40.8% | 41.2% |
| % obesity, prior to AMD | UKBB | 8.9% | 7.3% | 10.1% | 10.0% | 9.9% | 10.0% |
| % obesity, prior to AMD | eMERGE | 29.9% | 32.8% | 27.1% | 27.4% | 26.9% | 27.6% |
| % obesity, prior to AMD | MGBB | 33.1% | 36.0% | 33.1% | 33.2% | 32.8% | 33.3% |
| % obesity, prior to AMD | IAMDGC | NA | NA | NA | NA | NA | NA |
| % diabetes, prior to AMD | UKBB | 9.7% | 7.8% | 8.2% | 8.2% | 8.2% | 8.2% |
| % diabetes, prior to AMD | eMERGE | 20.1% | 19.5% | 20.4% | 20.4% | 20.1% | 20.5% |
| % diabetes, prior to AMD | MGBB | 28.5% | 28.7% | 17.9% | 18.6% | 19.2% | 18.4% |
| % diabetes, prior to AMD | IAMDGC | NA | NA | NA | NA | NA | NA |
| % hypertension, prior to AMD | UKBB | 47.0% | 43.2% | 39.4% | 39.5% | 39.3% | 39.6% |
| % hypertension, prior to AMD | eMERGE | 64.8% | 67.2% | 56.7% | 57.4% | 56.4% | 57.7% |
| % hypertension, prior to AMD | MGBB | 43.4% | 43.8% | 46.7% | 47.2% | 48.9% | 47.4% |
| % hypertension, prior to AMD | IAMDGC | NA | NA | NA | NA | NA | NA |
| % CAD, prior to AMD | UKBB | 7.9% | 6.9% | 7.0% | 7.0% | 6.9% | 7.0% |
| % CAD, prior to AMD | eMERGE | 8.8% | 8.7% | 8.7% | 8.7% | 8.3% | 8.9% |
| % CAD, prior to AMD | MGBB | 7.2% | 6.8% | 5.9% | 5.9% | 5.7% | 6.0% |
| % CAD, prior to AMD | IAMDGC | NA | NA | NA | NA | NA | NA |
| % tobacco use disorder, prior to AMD | UKBB | 20.8% | 14.8% | 18.9% | 18.8% | 18.7% | 18.9% |
| % tobacco use disorder, prior to AMD | eMERGE | 1.2% | 0.7% | 4.9% | 4.7% | 4.5% | 4.7% |
| % tobacco use disorder, prior to AMD | MGBB | 12.6% | 13.4% | 35.9% | 34.5% | 34.1% | 34.7% |
| % tobacco use disorder, prior to AMD | IAMDGC | NA | NA | NA | NA | NA | NA |
| % cataract, prior to AMD | UKBB | 24.8% | 26.7% | 13.5% | 13.8% | 13.7% | 13.9% |
| % cataract, prior to AMD | eMERGE | 39.7% | 48.6% | 12.9% | 15.0% | 14.7% | 15.2% |
| % cataract, prior to AMD | MGBB | 37.3% | 38.7% | 14.6% | 16.0% | 15.9% | 16.1% |
| % cataract, prior to AMD | IAMDGC | NA | NA | NA | NA | NA | NA |
| % glaucoma, prior to AMD | UKBB | 6.7% | 7.0% | 3.9% | 4.0% | 3.9% | 4.0% |
| % glaucoma, prior to AMD | eMERGE | 16.4% | 18.8% | 5.4% | 6.2% | 6.1% | 6.2% |
| % glaucoma, prior to AMD | MGBB | 13.8% | 14.9% | 6.3% | 6.8% | 6.7% | 6.8% |
| % glaucoma, prior to AMD | IAMDGC | NA | NA | NA | NA | NA | NA |
| % uveitis, prior to AMD | UKBB | 0.9% | 1.1% | 0.5% | 0.5% | 0.5% | 0.6% |
| % uveitis, prior to AMD | eMERGE | 3.4% | 3.5% | 0.8% | 0.9% | 0.9% | 0.9% |
| % uveitis, prior to AMD | MGBB | 3.30% | 4.0% | 1.1% | 1.3% | 1.4% | 1.3% |
| % uveitis, prior to AMD | IAMDGC | NA | NA | NA | NA | NA | NA |
