## Supplementary material for "Large-scale genetic analysis of age-related macular degeneration (AMD) drug targets highlights precision therapy opportunities for patients with high polygenic risk": Table S4

| Cohort | Pharmacomimetic score | Interactor | Interaction Beta | Interaction P-value | Interaction Beta (percent of AMD complement PRS interaction beta) |
|---|---|---|---|---|---|
| UKBB | C3 score | AMD complement PRS | 0.0684 | 2.9E-12 | 100% |
| UKBB | C3 score | CFH score | 0.0478 | 2.1E-06 | 70% |
| UKBB | C3 score | HTRA1 score | 0.0311 | 5.9E-04 | 45% |
| UKBB | C3 score | PRS without CFH or HTRA1 | 0.0399 | 8.7E-05 | 58% |
| UKBB | CFB score | AMD complement PRS | 0.0524 | 8.3E-07 | 100% |
| UKBB | CFB score | CFH score | 0.0370 | 7.2E-04 | 70% |
| UKBB | CFB score | HTRA1 score | 0.0180 | 7.4E-02 | 34% |
| UKBB | CFB score | PRS without CFH or HTRA1 | 0.0456 | 1.2E-05 | 87% |
| IAMDGC | C3 score | AMD complement PRS | 0.0722 | 1.5E-05 | 100% |
| IAMDGC | C3 score | CFH score | 0.0485 | 1.1E-03 | 67% |
| IAMDGC | C3 score | HTRA1 score | -0.0035 | 8.1E-01 | -5% |
| IAMDGC | C3 score | PRS without CFH or HTRA1 | 0.0049 | 7.3E-01 | 7% |
| IAMDGC | CFB score | AMD complement PRS | 0.0854 | 3.7E-08 | 100% |
| IAMDGC | CFB score | CFH score | 0.0701 | 9.1E-07 | 82% |
| IAMDGC | CFB score | HTRA1 score | 0.0437 | 2.7E-03 | 51% |
| IAMDGC | CFB score | PRS without CFH or HTRA1 | 0.0105 | 4.5E-01 | 12% |
| eMERGE | C3 score | AMD complement PRS | 0.0788 | 1.5E-05 | 100% |
| eMERGE | C3 score | CFH score | 0.0633 | 5.7E-04 | 80% |
| eMERGE | C3 score | HTRA1 score | 0.0365 | 3.6E-02 | 46% |
| eMERGE | C3 score | PRS without CFH or HTRA1 | 0.0282 | 1.2E-01 | 36% |
| eMERGE | CFB score | AMD complement PRS | 0.0407 | 3.6E-02 | 100% |
| eMERGE | CFB score | CFH score | 0.0627 | 1.5E-03 | 154% |
| eMERGE | CFB score | HTRA1 score | -0.0090 | 6.3E-01 | -22% |
| eMERGE | CFB score | PRS without CFH or HTRA1 | 0.0070 | 7.0E-01 | 17% |
| MGBB | C3 score | AMD complement PRS | 0.0286 | 1.7E-01 | 100% |
| MGBB | C3 score | CFH score | 0.0053 | 8.0E-01 | 19% |
| MGBB | C3 score | HTRA1 score | 0.0658 | 5.0E-02 | 230% |
| MGBB | C3 score | PRS without CFH or HTRA1 | 0.0150 | 4.9E-01 | 52% |
| MGBB | CFB score | AMD complement PRS | 0.0592 | 9.0E-03 | 100% |
| MGBB | CFB score | CFH score | 0.0645 | 5.0E-03 | 109% |
| MGBB | CFB score | HTRA1 score | 0.0256 | 4.9E-01 | 43% |

| | | | | | |
|---|---|---|---|---|---|
| MGBB | CFB score | PRS without CFH or HTRA1 | 0.0049 | 8.3E-01 | 8% |
