## Supplementary material for "Large-scale genetic analysis of age-related macular degeneration (AMD) drug targets highlights precision therapy opportunities for patients with high polygenic risk": Table S5

| UK Biobank OCT-derived Phenotype | Beta for Dry AMD association | SE for Dry AMD association | P-value for Dry AMD association | # Dry AMD Cases | # Dry AMD Controls |
|---|---|---|---|---|---|
| oct_isos_rpe_thickness_central_subfield_sd | -0.256 | 0.030 | 2.90E-17 | 744 | 20066 |
| oct_isos_rpe_thickness_inner_subfield_sd | -0.242 | 0.029 | 1.35E-16 | 743 | 20055 |
| oct_isos_rpe_thickness_sd | -0.221 | 0.029 | 1.34E-14 | 741 | 20020 |
| oct_isos_rpe_thickness_outer_subfield_sd | -0.212 | 0.029 | 2.34E-13 | 741 | 20020 |
| oct_inl_rpe_thickness_central_subfield_sd | -0.149 | 0.029 | 3.04E-07 | 744 | 20066 |
| oct_gcipl_thickness_sd | -0.185 | 0.039 | 2.43E-06 | 741 | 20020 |
| oct_macular_thickness_inner_temporal_subfield_sd | -0.142 | 0.031 | 5.03E-06 | 724 | 19849 |
| oct_macular_thickness_inner_inferior_subfield_sd | -0.139 | 0.031 | 6.40E-06 | 723 | 19838 |
| oct_rpe_thickness_central_subfield_sd | 0.273 | 0.061 | 7.74E-06 | 268 | 9410 |
| oct_elm_isos_thickness_inner_subfield_sd | 0.160 | 0.039 | 4.45E-05 | 743 | 20055 |
| oct_macular_thickness_inner_nasal_subfield_sd | -0.125 | 0.031 | 5.42E-05 | 724 | 19849 |
| oct_macular_thickness_inner_superior_subfield_sd | -0.127 | 0.032 | 6.80E-05 | 724 | 19849 |
| oct_elm_isos_thickness_sd | 0.153 | 0.040 | 1.19E-04 | 741 | 20020 |
| oct_rpe_thickness_inner_inferior_subfield_sd | 0.224 | 0.059 | 1.33E-04 | 268 | 9410 |
| oct_elm_isos_thickness_outer_subfield_sd | 0.143 | 0.039 | 2.78E-04 | 741 | 20020 |
| oct_inl_rpe_thickness_inner_subfield_sd | -0.104 | 0.030 | 5.01E-04 | 743 | 20055 |
| oct_inl_elm_thickness_central_subfield_sd | -0.113 | 0.034 | 7.63E-04 | 744 | 20066 |
| oct_macular_thickness_sd | -0.103 | 0.032 | 1.36E-03 | 741 | 20020 |
| oct_macular_thickness_outer_inferior_subfield_sd | -0.101 | 0.038 | 7.11E-03 | 723 | 19834 |
| oct_rpe_thickness_inner_nasal_subfield_sd | 0.154 | 0.060 | 1.05E-02 | 268 | 9410 |
| oct_inl_rpe_thickness_sd | -0.075 | 0.032 | 1.73E-02 | 741 | 20020 |
| oct_macular_thickness_outer_temporal_subfield_sd | -0.085 | 0.036 | 1.77E-02 | 724 | 19846 |
| oct_rpe_thickness_inner_temporal_subfield_sd | 0.139 | 0.059 | 1.87E-02 | 268 | 9410 |
| oct_macular_thickness_outer_superior_subfield_sd | -0.075 | 0.036 | 3.55E-02 | 724 | 19845 |
| oct_inl_thickness_sd | -0.075 | 0.036 | 3.81E-02 | 741 | 20020 |
| oct_rpe_thickness_sd | 0.060 | 0.029 | 4.08E-02 | 741 | 20020 |
| oct_elm_isos_thickness_central_subfield_sd | 0.074 | 0.038 | 5.49E-02 | 744 | 20066 |
| oct_inl_rpe_thickness_outer_subfield_sd | -0.060 | 0.033 | 6.62E-02 | 741 | 20020 |
| oct_rpe_thickness_inner_superior_subfield_sd | 0.099 | 0.061 | 1.05E-01 | 268 | 9410 |
| oct_inl_elm_thickness_inner_subfield_sd | -0.055 | 0.035 | 1.21E-01 | 743 | 20055 |

| | | | | | |
|---|---|---|---|---|---|
| oct_macular_thickness_central_subfield_sd | -0.057 | 0.037 | 1.23E-01 | 724 | 19849 |
| oct_macular_thickness_outer_nasal_subfield_sd | -0.051 | 0.036 | 1.61E-01 | 724 | 19849 |
| oct_rpe_thickness_outer_inferior_subfield_sd | 0.076 | 0.061 | 2.07E-01 | 268 | 9410 |
| oct_rnfl_thickness_sd | 0.037 | 0.032 | 2.45E-01 | 741 | 20020 |
| oct_rpe_thickness_outer_superior_subfield_sd | 0.050 | 0.061 | 4.14E-01 | 268 | 9410 |
| oct_inl_elm_thickness_outer_subfield_sd | 0.020 | 0.036 | 5.89E-01 | 741 | 20020 |
| oct_vcdr_sd | -0.015 | 0.040 | 7.05E-01 | 672 | 18611 |
| oct_rpe_thickness_outer_nasal_subfield_sd | 0.013 | 0.064 | 8.40E-01 | 268 | 9410 |
| oct_rpe_thickness_outer_temporal_subfield_sd | 0.011 | 0.064 | 8.60E-01 | 268 | 9410 |
| oct_inl_elm_thickness_sd | -0.003 | 0.037 | 9.30E-01 | 741 | 20020 |
